# Lower Risks of Incident Colorectal Cancer in SGLT2i Users Compared to DPP4i Users: A Propensity Score-matched Study with Competing Risk Analysis

**DOI:** 10.1101/2022.07.16.22277673

**Authors:** Raymond Ngai Chiu Chan, Robert Ngai Fung Chan, Oscar Hou In Chou, Teddy Tai Loy Lee, Leonardo Roever, Guoliang Li, Wing Tak Wong, Abraham Ka Chung Wai, Tong Liu, Gary Tse, Sharen Lee

## Abstract

**Background:** Diabetes mellitus is associated with the development of colorectal cancer (CRC). There have been a lack of study comparing the risk of colorectal cancer in sodium-glucose co-transporter 2 inhibitors (SGLT2i) and dipeptidyl peptidase 4 inhibitors (DPP4i), both of which commonly prescribed second line agents for diabetes.

**Methods:** We conducted a territory wide retrospective cohort study on patients with type 2 diabetes who was prescribed either of the two agents. Baseline demographics, use of other medications, comorbidities and biochemical parameters were extracted. Propensity score matching was performed to reduce the impacts of cofounders. Cause specific Cox regression was used to evaluate the risk of incident colorectal cancer in SGLT2i users, as compared to DPP4i users. Subgroup analyses based on age, gender and estimated glomerular filtration rate were performed.

**Results:** After propensity score matching, we included 13029 subjects who were prescribed SGLT2i and DPP4i respectively. Incidence rate ratio of CRC was 0.566 (0.418-0.766) in SGLT2i users. Overall, use of SGLT2i was associated with a lower risk of incident CRC (HR: 0.526; 95% CI: 0.382-0.724; P <0.001). In subgroup analyses, use of SGLT2i was associated with lower risks of incident CRC only in men (HR: 0.461; 95% CI: 0.303-0.702; P <0.001), patients < 65 years old and patients (HR:0.294; 95% CI: 0.174-0.496; P<0.001) with eGFR ≥ 45 mL/min/ 1.73m^2^ (HR: 0.560; 95% CI: 0.395-0.792; P =0.001).

**Conclusion:** Use of SGLT2i may reduce risk of incident CRC as compared to use of DPP4i, especially in younger male patients with fairly preserved renal function.

## INTRODUCTION

Diabetes mellitus is a global public health issue affecting 536.6 million people worldwide in 2021 ^1^. Previous studies have established clear epidemiological links between diabetes mellitus and various co-morbidities, including malignancies like liver, pancreatic, endometrial and colorectal cancers (CRC) ^2, 3^. A meta-analysis in 2013, which pooled data from 20 controlled trials and cohort studies after 2007, reported a higher risk of incident CRC as well as a higher CRC-specific mortality in diabetic patients ^4^. Several groups have investigated the effects of anti-diabetic medications on the risk of CRC. Metformin, an oral anti-hyperglycemic agent recommended by the American Diabetes Association as the preferred initial pharmacologic treatment for type 2 diabetes mellitus (T2DM), was found to have a protective effect on incident CRC by several studies ^5-7^, although conflicting results have been generated by other groups ^8-11^. The effects of newer anti-diabetic medications, especially dipeptidyl peptidase 4 inhibitors (DPP4i) and sodium-glucose co-transporter 2 inhibitors (SGLT2i), were less studied.

DPP4i and SGLT2i are two emerging drug classes approved by the FDA for the treatment of T2DM ^12^. Given the protective effects of DPP4i on pancreatic beta cells function, as well as the cardioprotective and renoprotective effects of SGLT2i, both are increasingly prescribed as second line treatment for T2DM ^13-16^. Preclinical studies on DPP4i and SGLT2i have highlighted their potentials in CRC suppression ^17-19^. Although there are few studies evaluating the effects of DPP4i on CRC, most of them were of small scale and none directly compared it with SGLT2i ^20^. Whether any of these agents could alter the risk of incident CRC in T2DM patients remains unclear. Hence in the present study, we aim to compare the risk of incident CRC in DPP4i and SGLT2i users amongst T2DM patients using a large territory wide cohort in Hong Kong.

## METHODS

### Study design and population

This was a territory-wide retrospective cohort study in T2DM patients treated with SGLT2i or DPP4i between January 1st, 2016, and December 31st, 2019 in Hong Kong. Patients were followed up until December 31st, 2020, or death, whichever came earlier. The current study was approved by The Joint Chinese University of Hong Kong–New Territories East Cluster Clinical Research Ethics Committee. Subjects were identified from the Clinical Data Analysis and Reporting System (CDARS) of the Hospital Authority in Hong Kong. It is a territory-wide database that centralizes patient information from all public hospitals and clinics in Hong Kong, which facilitates retrieval of clinical characteristics, disease diagnosis, laboratory results and drug treatment details. It has been used by local teams in Hong Kong to conduct studies on T2DM ^21-23^, and recently by our team comparing the cardiovascular outcomes between SGLT2I and DPP4I users ^24-26^.

Patients were excluded if any of the following criteria were met: 1) less than one year of drug exposure; 2) on both DPP4I and SGLT2I, or switched between the two drug classes; 3) died within 30 days after initial drug exposure; 4) less than 18 years old at the start of the study; 5) pregnancy; 6) without complete demographics 7) with no HbA1c records at baseline; 8) with a history of colorectal cancer prior to baseline.

Patients’ demographics, clinical and biochemical data were extracted for the present study. Comorbidities at baseline were defined and extracted using the International Classification of Diseases Ninth Edition (ICD-9) codes (**Supplementary Table 1**). Charlson’s standard comorbidity index was calculated. Use of anti-diabetic and lipid-lowering agents were extracted. Baseline biochemistry, including the complete blood count, renal function tests, liver function tests, lipid and glycemic profiles were extracted. Estimated glomerular filtration rate (eGFR) was calculated by the abbreviated modification of diet in renal disease (MDRD) formula ^27^.

### Adverse outcomes and statistical analysis

The primary endpoint of the present study was incident colorectal cancer (ICD-9: 153-154). Mortality data were obtained from the Hong Kong Death Registry, a population-based official government registry with the registered death records of all Hong Kong citizens linked to CDARS. The endpoint date of interest for eligible patients was the event presentation date. The endpoint for those without primary outcome presentation was the mortality date or the endpoint of the study (December 31st, 2020).

For baseline clinical characteristics, continuous and categorical variables were presented as mean (standard deviation [SD]) and frequency (percentage) respectively. Propensity score matching (PSM) with a 1:1 ratio for SGLT2i users versus DPP4i users based on age, gender, Charlson’s comorbidity index, other comorbidities and non-SGLT2I/ DPP4I medications was performed using the nearest neighbor search strategy. Baseline characteristics between patients with SGLT2i and DPP4i use before and after matching were compared with standardized mean difference (SMD), with SMD<0.20 regarded as well-balanced. The incidence of new-onset colorectal cancer was derived from dividing the number of outcomes by person-year at risks, which estimate the number of years at risks. Cause specific hazard models were used to calculate the unadjusted and adjusted hazard ratios. Important covariates were adjusted by backward selection. HDL was considered as a surrogate marker of BMI and was adjusted in addition to other covariates. It has been shown to correlate better with BMI as compared to other serum lipids ^28-31^. Fine Gray’s subdistribution hazard models were conducted as sensitivity analyses. Subgroup analyses were performed in patients ≥ 65 years old, < 65 years old, male and female. We aimed to evaluate the impact of renal function on the effects of SGLT2i and/or DPP4i, hence subgroup analyses were also performed in patients with eGFR ≥ 45 mL/min/ 1.73m^2^ and <45 mL/min/ 1.73m^2^ respectively. The hazard ratios (HR), 95% CI and P-value were reported. Statistical significance was taken as P-value < 0.05. All statistical analyses were performed with RStudio software (Version: 1.1.456).

## RESULTS

### Baseline characteristics of the study population

In this retrospective cohort study, 86 353 patients in Hong Kong using SGLT2i/DPP4i between January 1^st^, 2016 and December 31^st^, 2019 were recruited. They were followed up until December 31^st^, 2020 or deaths, which ever came earlier. Patients with less than one month of drug exposure (n = 3019), on both DPP4i and SGLT2i (n = 13529), died within 30 days after initial drug exposure (n = 4491), less than 18 years old at baseline (n = 592), less than 1 year of drug exposure (n = 1781), pregnancy (n = 19), without complete demographics or mortality data (n = 15), without baseline HbA1c (n = 9766), with CRC prior to baseline (n = 289) were excluded.

Subsequently, 18 741 SGLT2i users and 33 839 DPP4i users were included. After 1:1 propensity score matching, 13 029 patients on SGLT2i and DPP4i respectively entered statistical analyses. In the matched cohort, 65 (0.5%) SGLT2i users and 118 (0.9%) DPP4i users developed incident CRC. The baseline characteristic of patients before and after PSM are shown in **Supplementary Table 2 and Table 1** respectively.

### SGLT2i use is associated with reduced risk of incident CRC

Over a follow-up period of 117734.1 person-years, 183 incident CRC were identified. Overall, the incidence of CRC (IRR: 0.566; 95% CI: 0.418-0.766; P<0.001) were lower amongst SGLT2i users compared to DPP4i users after PSM **(Table 2)**. We used univariable Cox regression to identify the potential risk factors of incident CRC before and after PSM, which were serially adjusted in subsequent multivariable models **(Supplementary Table 3 and 4)**. Unadjusted and adjusted hazard ratios for incident CRC were presented in **Table 3**. In all models, use of SGLT2i was significantly associated with lower risks of incident CRC as compared to use of DPP4i. After adjusting for age, gender, HbA1c, use of other medications, comorbidities and HDL, use of SGLT2i was associated with an approximately 47.4% reduction in the risk of incident CRC (HR: 0.526; 95% CI: 0.382-0.724; P<0.001).

**Table 1:**
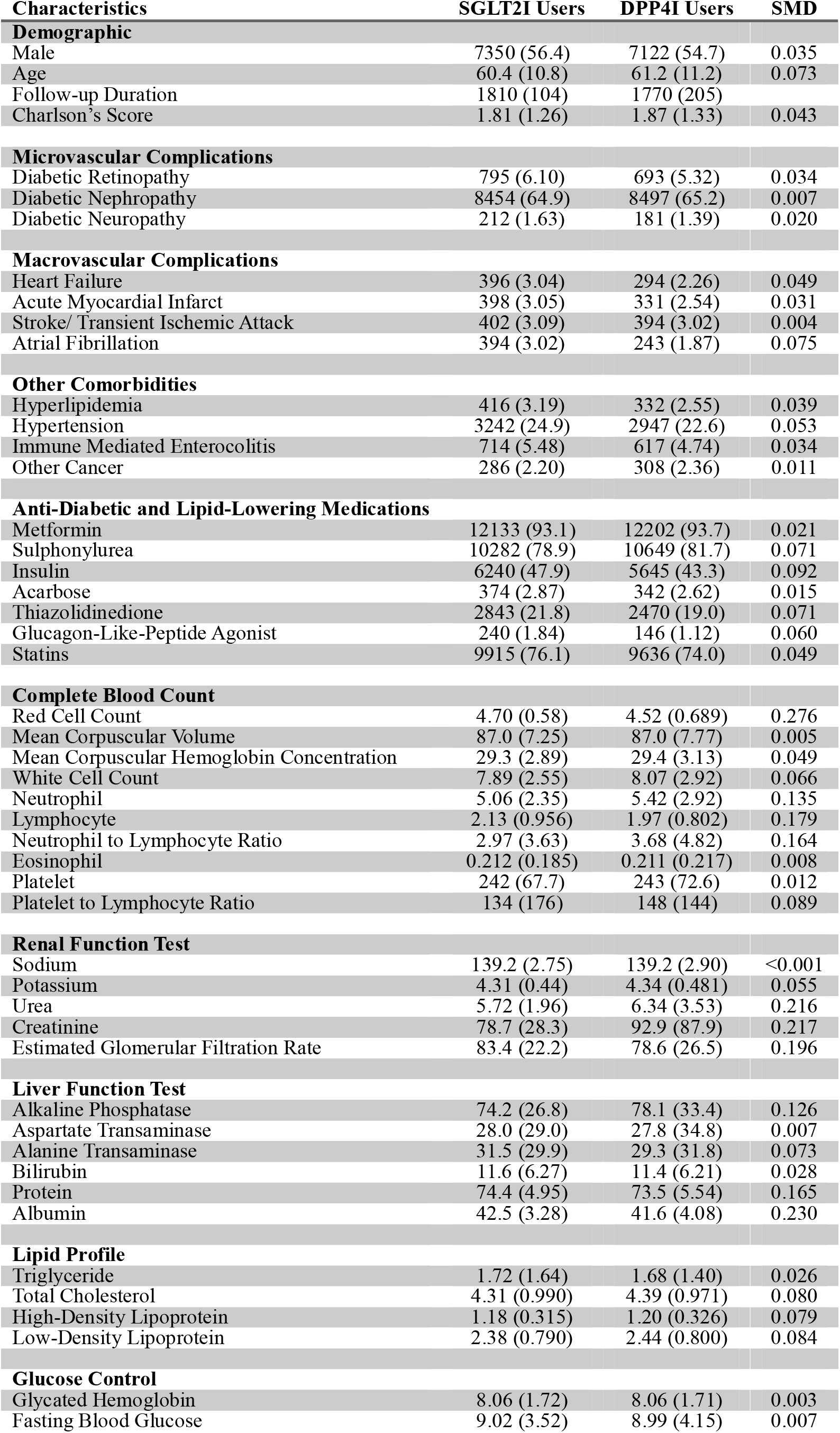
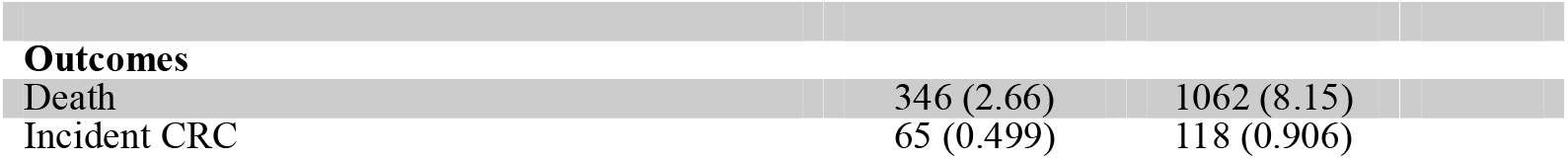
Baseline Demographics of SGLT2i and DPP4i Users after Propensity Score Matching

**Table 2:** Annualized Incidence Rate of CRC per 1000 Person-Year before and after Propensity Score Matching.

|  | **Person-Years** | **Number of Event** | **IR [95% CI]** | **IRR [95% CI]** |
| --- | --- | --- | --- | --- |
| **Before Propensity Score Matching** |  |  |  |  |
| **Overall** | 252767 | 471 | 1.86 [1.70-2.04] | - |
| **DPP4i Users** | 159796.7 | 377 | 2.36 [2.13-2.61] | - |
| **SGLT2i Users** | 92970.32 | 94 | 1.01 [0.817-1.24] | 0.429 [0.342-0.537] |
| **After Propensity Score Matching** |  |  |  |  |
| **Overall** | 117734.1 | 183 | 1.55 [1.34- 1.80] | - |
| **DPP4i Users** | 58083.66 | 118 | 2.03 [1.68-2.43] | - |
| **SGLT2i Users** | 59650.41 | 65 | 1.09 [0.841-1.39] | 0.566 [0.418-0.766] |

**Table 3:** Univariable and Multivariable Cause-Specific Cox Regression after Propensity Score Matching Model 1: Unadjusted hazard ratio. Model 2: Adjusted for age and gender. Model 3: Adjusted for age, gender and HbA1c. Model 4: Adjusted for age, gender, HbA1c and use of other medications. Model 5: Adjusted for age, gender, HbA1c, use of medications and comorbidities. Model 6: Adjusted for age, gender, HbA1c, use of medications, comorbidities, and HDL.

|  | **Hazard ratios** | **95% CI** | **P-value** |
| --- | --- | --- | --- |
| **Before Propensity Score Matching** |  |  |  |
| **Model 1** | 0.424 | 0.338-0.531 | <0.001 |
| **Model 2** | 0.599 | 0.474-0.760 | <0.001 |
| **Model 3** | 0.599 | 0.474-0.759 | <0.001 |
| **Model 4** | 0.579 | 0.444-0.758 | <0.001 |
| **Model 5** | 0.588 | 0.449-0.769 | <0.001 |
| **Model 6** | 0.572 | 0.431-0.759 | 0.001 |
| **After Propensity Score Matching** |  |  |  |
| **Model 1** | 0.531 | 0.392-0.719 | <0.001 |
| **Model 2** | 0.548 | 0.405-0.742 | <0.001 |
| **Model 3** | 0.548 | 0.405-0.742 | <0.001 |
| **Model 4** | 0.537 | 0.397-0.728 | <0.001 |
| **Model 5** | 0.535 | 0.395-0.725 | <0.001 |
| **Model 6** | 0.526 | 0.382-0.724 | <0.001 |

Sensitivity analyses were performed using Fine-Gray’s subdistribution regression model to confirm the predictiveness of the models. Similarly, use of SGLT2i was associated with lower risks of incident CRC in the sensitivity analyses after serial adjustment of important covariates identified in univariable analyses (**Supplementary Table 5**). In model 5, use of SGLT2i was associated with a 42.5% reduction in risk of incident CRC (HR: 0.575; 95% CI: 0.368-0.896; P= 0.015).

### Effects of SGLT2i on the risk of incident CRC stratified by age and gender

Sub-group analyses based on age and gender were further performed. In male, use of SGLT2i was associated with lower risks of incident CRC (HR: 0.461; 95% CI: 0.303-0.702; P <0.001) but no significant association was found in female **(Table 4)**. Similar results were reproduced in the sensitivity analyses using Fine-Gray’s subdistribution hazard models (HR: 0.505; 95% CI: 0.339-0.752; P <0.001) **(Supplementary Table 6)**. In patients younger than 65 years old, use of SGLT2i was associated with reduced risks of incident CRC (HR: 0.294; 95% CI: 0.174-0.496; P <0.001) whereas no significant association was found in the older group **(Table 5)**. Again, similar results were observed in sensitivity analyses (HR: 0.328; 95% CI: 0.198-0.546; P<0.001). **(Supplementary Table 7)**

**Table 4:** Univariable and Multivariable Cause-Specific Cox Regression after Propensity Score Matching in Patient Subgroup Stratified by Gender Model 1: Unadjusted hazard ratio. Model 2: Adjusted for age. Model 3: Adjusted for age and HbA1c. Model 4: Adjusted for age, HbA1c and use of other medications. Model 5: Adjusted for age, HbA1c, use of medications and comorbidities. Model 6: Adjusted for age, HbA1c, use of other medications, comorbidities and HDL.

|  | Hazard ratios | 95% CI | P-value |
| --- | --- | --- | --- |
| **Male** |  |  |  |
| **Model 1** | 0.467 | 0.314-0.692 | <0.001 |
| **Model 2** | 0.481 | 0.324-0.714 | <0.001 |
| **Model 3** | 0.480 | 0.323-0.713 | <0.001 |
| **Model 4** | 0.481 | 0.324-0.715 | <0.001 |
| **Model 5** | 0.478 | 0.322-0.710 | <0.001 |
| **Model 6** | 0.461 | 0.303-0.702 | <0.001 |
| **Female** |  |  |  |
| **Model 1** | 0.639 | 0.398-1.03 | 0.064 |
| **Model 2** | 0.481 | 0.324-0.714 | 0.097 |
| **Model 3** | 0.670 | 0.417-1.08 | 0.098 |
| **Model 4** | 0.624 | 0.388-1.01 | 0.053 |
| **Model 5** | 0.624 | 0.387-1.01 | 0.053 |
| **Model 6** | 0.616 | 0.374-1.01 | 0.056 |

**Table 5:**
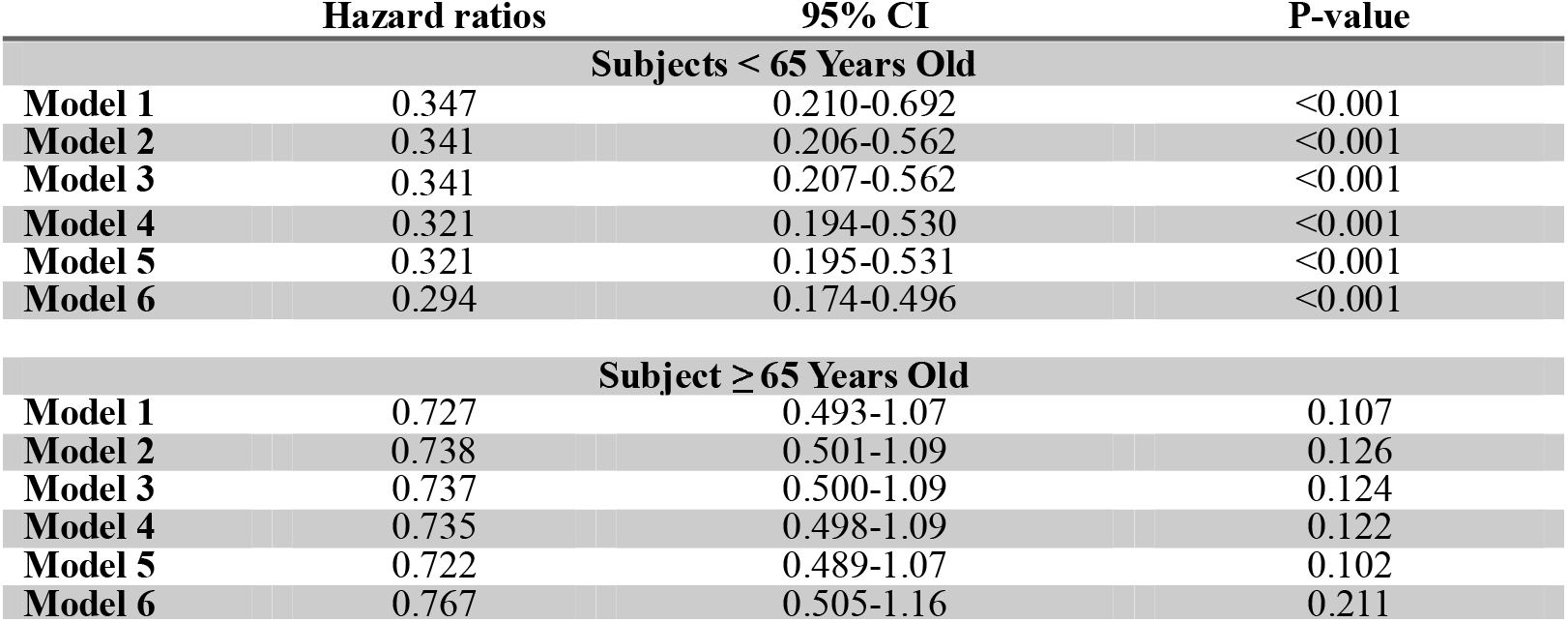
Univariable and Multivariable Cause-Specific Cox Regression after Propensity Score Matching in Patient Subgroup Stratified by Age Model 1: Unadjusted hazard ratio. Model 2: Adjusted for age and gender. Model 3: Adjusted for age, gender and HbA1c. Model 4: Adjusted for age, gender, HbA1c and use of other medications. Model 5: Adjusted for age, gender, HbA1c, use of medications and comorbidities. Model 6: Adjusted for age, gender, HbA1c, use of other medications, comorbidities and HDL.

|  | Hazard ratios | 95% CI | P-value |
| --- | --- | --- | --- |
| **Subjects < 65 Years Old** |  |  |  |
| **Model 1** | 0.347 | 0.210-0.692 | <0.001 |
| **Model 2** | 0.341 | 0.206-0.562 | <0.001 |
| **Model 3** | 0.341 | 0.207-0.562 | <0.001 |
| **Model 4** | 0.321 | 0.194-0.530 | <0.001 |
| **Model 5** | 0.321 | 0.195-0.531 | <0.001 |
| **Model 6** | 0.294 | 0.174-0.496 | <0.001 |
| **Subject ≥ 65 Years Old** |  |  |  |
| **Model 1** | 0.727 | 0.493-1.07 | 0.107 |
| **Model 2** | 0.738 | 0.501-1.09 | 0.126 |
| **Model 3** | 0.737 | 0.500-1.09 | 0.124 |
| **Model 4** | 0.735 | 0.498-1.09 | 0.122 |
| **Model 5** | 0.722 | 0.489-1.07 | 0.102 |
| **Model 6** | 0.767 | 0.505-1.16 | 0.211 |

### Effects of SGLT2i on the risk of incident CRC stratified by eGFR

To assess the impact of eGFR on the effects of SGLT2i, we conducted subgroup analyses based on the eGFR. As the number of incident CRC in patients with eGFR< 45 mL/min/ 1.73m^2^ is inadequate to perform a robust analysis, the unmatched cohort were used. In patients with eGFR ≥ 45 mL/min/ 1.73m^2^, use of SGLT2i was correlated with a significantly reduced risk of incident CRC (HR: 0.560; 95% CI: 0.395-0.792; P = 0.001). However, in patients with more advanced CKD, the association was lost. (**Table 6)** Sensitivity analyses yielded similar findings (**Supplementary Table 8)**.

**Table 6:**
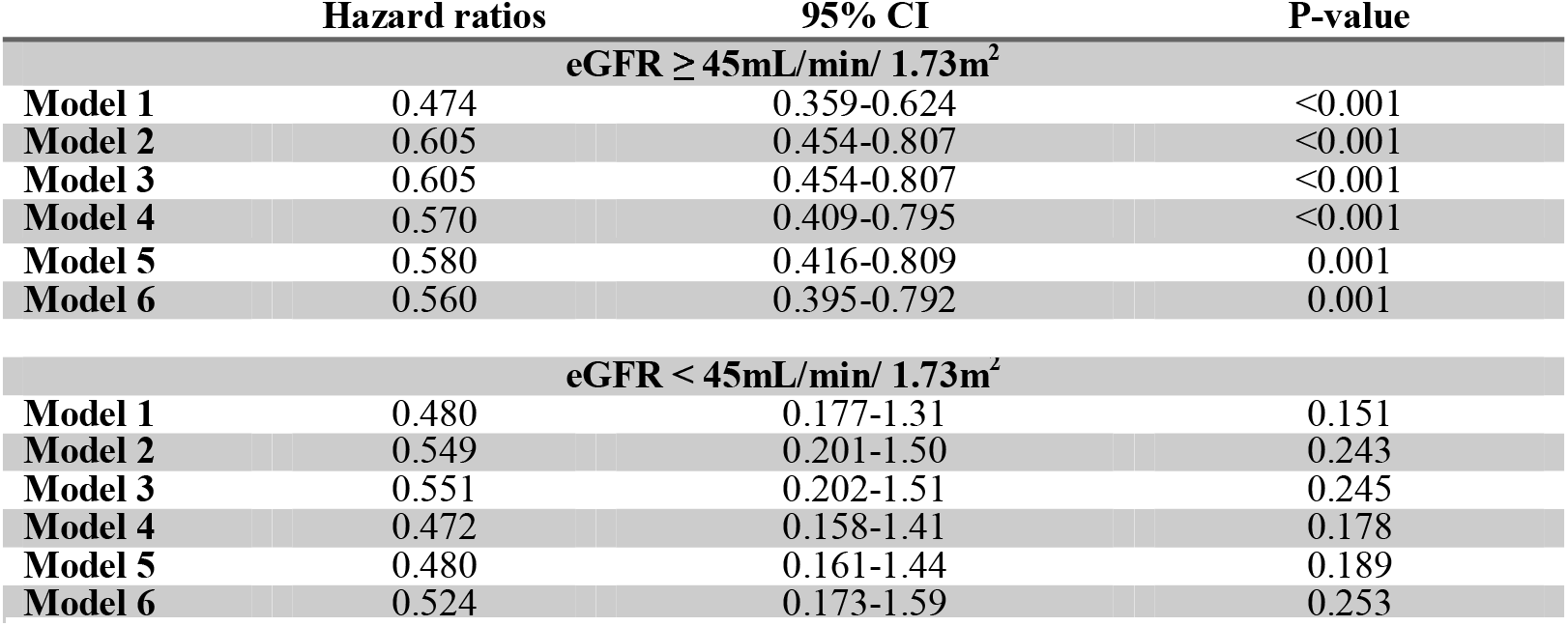
Univariable and Multivariable Cause-Specific Cox Regression before Propensity Score Matching in Patient Subgroup Stratified by eGFR Model 1: Unadjusted hazard ratio. Model 2: Adjusted for age and gender. Model 3: Adjusted for age, gender and HbA1c. Model 4: Adjusted for age, gender, HbA1c and use of other medications. Model 5: Adjusted for age, gender, HbA1c, use of medications and comorbidities. Model 6: Adjusted for age, gender, HbA1c, use of medications, comorbidities and HDL.

|  | Hazard ratios | 95% CI | P-value |
| --- | --- | --- | --- |
| **eGFR ≥ 45mL/min/ 1.73m<sup>2</sup>** |  |  |  |
| **Model 1** | 0.474 | 0.359-0.624 | <0.001 |
| **Model 2** | 0.605 | 0.454-0.807 | <0.001 |
| **Model 3** | 0.605 | 0.454-0.807 | <0.001 |
| **Model 4** | 0.570 | 0.409-0.795 | <0.001 |
| **Model 5** | 0.580 | 0.416-0.809 | 0.001 |
| **Model 6** | 0.560 | 0.395-0.792 | 0.001 |
| **eGFR < 45mL/min/ 1.73m<sup>2</sup>** |  |  |  |
| **Model 1** | 0.480 | 0.177-1.31 | 0.151 |
| **Model 2** | 0.549 | 0.201-1.50 | 0.243 |
| **Model 3** | 0.551 | 0.202-1.51 | 0.245 |
| **Model 4** | 0.472 | 0.158-1.41 | 0.178 |
| **Model 5** | 0.480 | 0.161-1.44 | 0.189 |
| **Model 6** | 0.524 | 0.173-1.59 | 0.253 |

### SGLT2i use is associated with lower all-cause mortality in patients who developed incident CRC

We compared the all-cause mortality of patients with incident CRC amongst SGLT2i and DPP4i users. Given the limited number of deaths with incident CRC in the matched cohort, the analyses were, again, performed in the unmatched cohort. Use of SGLT2i was not associated with lower risks of all-cause mortality (HR: 0.285; 95% CI: 0.066-1.25; P = 0.097) after adjusted for age, gender HbA1c, use of medications, and comorbidities **(Supplementary Table 9)**.

## DISCUSSION

In the present study, we used a territory-wide cohort to compare the risk of incident CRC amongst SGLT2i and DPP4i users. To our knowledge, this is the first study thus far to evaluate the incidence of CRC amongst patients treated with the named medications. Importantly, we were able to demonstrate several clinically relevant findings: 1) use of SGLT2i was associated with a lower risk of incident CRC after serial adjustment. 2) The effects of SGLT2i on incident CRC, as compared to DPP4i, differ with respect to patients’ age, gender and renal function. 3) Use of SGLT2i may not reduce all-cause mortality in DM patients with incident CRC. These findings are of clinical significance as SGLT2i and DPP4i are both commonly prescribed second line pharmacological treatment for T2DM.

### SGLT2i Reduce Risk of Incident CRC via an eGFR-Dependent Pathway

Diabetes mellitus and insulin resistance have been recognized to be major risk factors of various malignancies including CRC, although the exact mechanism remains unclear ^2, 3^. Hyperinsulinemia, which characterizes pre-diabetes and early diabetes, was known to promote cancer cell survival and mitogenesis via binding to insulin and insulin-like growth factor (IGF) receptors that are widely expressed on cancer cells membrane ^32-34^. Other mechanisms, such as upregulation of inflammatory cytokines, alterations of cellular energetics and shared risk factors, have previously been proposed ^2, 35^. In view of the epidemiological and biological links between CRC and diabetes mellitus, studies have investigated the relationship between different anti-diabetic medications and incident CRC.

Currently, there is a lack of evidence on the effects of SGLT2i, a relatively new agent for T2DM that inhibits SGLT2 in the kidney to promote glycosuria, on incident CRC ^36^. In contrary to most anti-diabetic medications, its major mechanism of action is independent of insulin. Studies suggested that use of SGLT2i improved insulin sensitivity and potentially lowers plasma insulin level ^37, 38^. Preclinical animal models echoed this finding by showing a dramatically reduced plasma insulin in SGLT2 knockout mice ^39^. Furthermore, use of SGLT2i has been shown to improve shared risk factors between CRC and diabetes, reduce circulating inflammatory cytokines, attenuate vascular endothelial and smooth muscle cells proliferation in response to interleukins, alter cellular energetics and suppress oxidative stress, all of which may be protective factors against malignancies such as CRC ^37, 40-42^. However, whether SGLT2i reduces risk of incident CRC and the actual mechanisms, if any, remain unknown.

In the present study, we demonstrated that SGLT2i significantly reduced the risk of incident CRC, and the association was only significant when the eGFR was ≥ 45 mL/min/ 1.73m^2^. Apparently, the glycosuria effects of SGLT2i depends on relatively preserved glomerular function. A previous study suggested that the glucose lowering effect of SGLT2i was attenuated when the eGFR dropped to < 45 mL/min/ 1.73m^2^, while another suggested that the effect was lost when the eGFR dropped to <30 mL/min/ 1.73m^2^ ^43, 44^. In our study, we were able to demonstrate that the protective effect of SGLT2i against incident CRC, as compared to DPP4i, was independent of glycemic control by adjusting for HbA1c, but the association was lost when the eGFR was below 45 mL/min/ 1.73m^2^. Hence, it is reasonable to hypothesize that SGLT2i may reduce the risk of incident CRC via its glycosuria effect, which reduces circulating insulin level at any given blood glucose level and downregulates farnesylatesd Ras protein to supress mitogenesis ^45^. Although the role of chronic inflammation and SGLT2 expression on CRC cell lines has been previously implicated, a recent study showed that oral administration of SGLT2i in obese and diabetic mice with azoxymethane-induced colorectal pre-neoplastic lesions appeared to reduce IGF-1 and several related signaling molecules in the colonic mucosa, while direct administration of SGLT2i on human CRC cell lines exhibited no impacts on cellular proliferation, further supporting our hypothesis ^46^. Further investigations are warranted to validate these findings.

### Differential effects of SGLT2i across gender and age

In the present study, we also demonstrated that SGLT2i only reduced risk of incident CRC in male and younger patients, but not their female or older counterparts. In fact, gender dimorphism in CRC in terms of the incidence, molecular pathogenesis and prognosis have been previously reported ^47, 48^. Lifestyle factors, hormonal differences, and more recently single nucleotide polymorphisms or genetic variants have been proposed to explain these differences ^49-52^.

Several epidemiological studies have suggested that the relationship between obesity and CRC is stronger in male than female, potentially due to the fact the protective effect of estrogen over insulin resistance and subsequent hyperinsulinemia ^53-57^. Recent studies also suggested the insulin-IGF axis is preferentially upregulated in men with CRC ^58, 59^. Differences in the level and biological action of insulin and IGF across the two genders may potentially account for the discrepancy. Meanwhile, elderly patients with T2DM may have relatively less preserved pancreatic β-cell function and renal function while insulin sensitivity appeared to be similar in older and younger subjects with comparable BMI ^60-62^. These may limit the effects of SGLT2i on preventing incident CRC.

### Strengths and limitations

With the use of medical records from a territory-wide database, CDARS, our study was able to detect relatively rare outcomes, such as incident CRC, with an adequate sample size and follow-up duration. The present study was also the first to compare the difference in CRC incidence amongst T2DM patients treated with SGLT2i and DPP4i. Subgroup analyses in our study may also provide mechanistic insight into the anti-tumour effects of SGLT2i. However, there are still certain limitation in our study. First, although CDARS has been considered a relatively reliable source of clinical data, there was unavoidably information bias due to the risk of under-coding, coding errors and missing data. Second, BMI has been considered an important risk factor of CRC but was not available in our database. The use of HDL as a surrogate marker may potentially and partially adjust for this limitation. Third, the retrospective nature of our study did not allow measurement of drug concentration, which might correlate with the outcomes. Fourth, the present study aimed at assessing the epidemiological links between use of SGLT2i and DPP4i with incident CRC and could not address the causal or mechanistic relationship. Lastly, SGLT2I and DPP4I are drug classes constituting a variety of agents from different brands with their respective formula. The present study did not conduct further analyses to compare the effects of different agents in the same drug class.

## CONCLUSION

SGLT2i use is associated with a reduced risk of incident CRC compared to DPP4i use. The association was significant only in younger patients, male and in patients with an eGFR ≥ 45 mL/min/ 1.73m^2^. SGLT2i use was not associated with a lower all-cause mortality in diabetic patients with incident CRC. Further studies are warranted to validate the findings.

## Data Availability

All data produced in the present study are available upon reasonable request to the authors

## ACKNOWLEDGEMENTS

Nil

---

**Figure 1:**
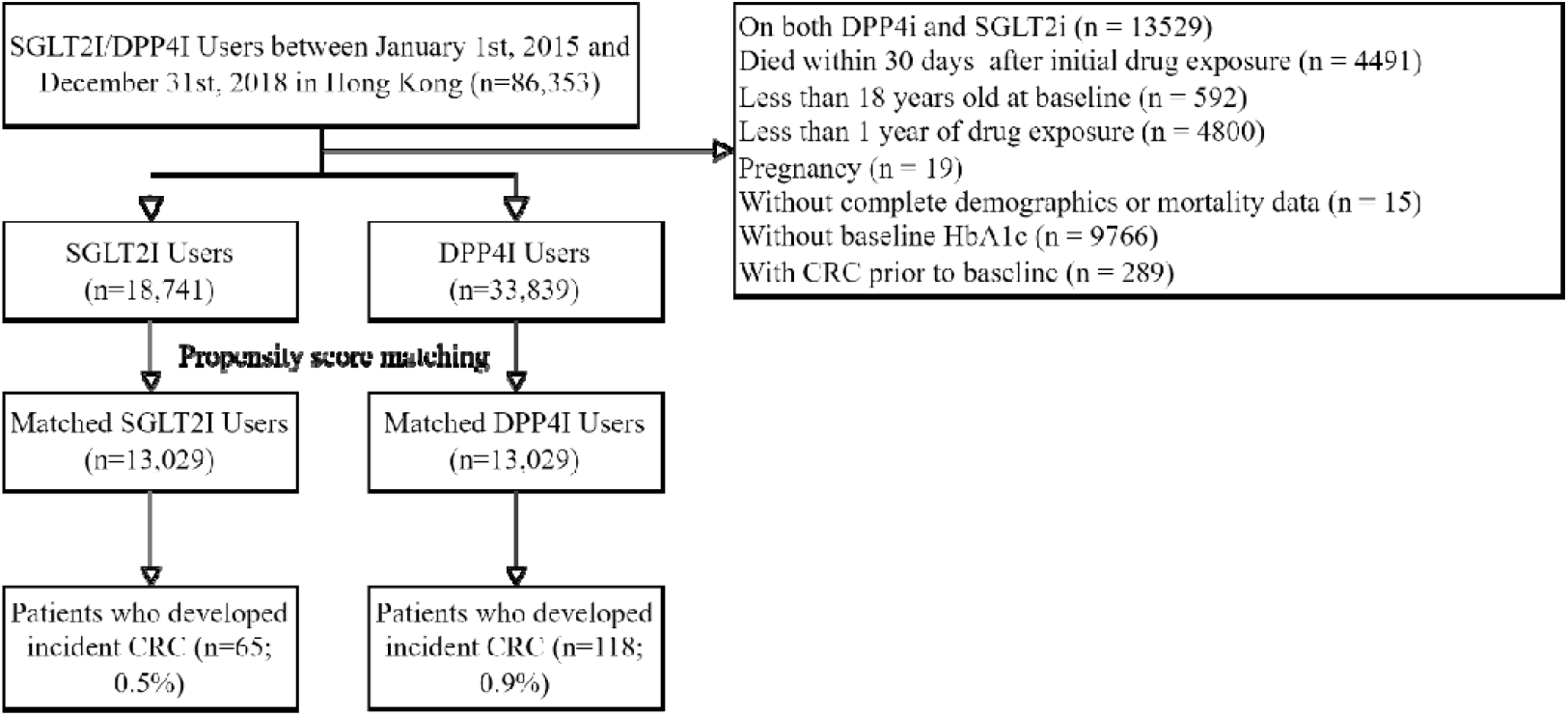
Flow Diaphragm for Subjects Selection and Propensity Score Matching

