## Supplementary material for "Lower Risks of Incident Colorectal Cancer in SGLT2i Users Compared to DPP4i Users: A Propensity Score-matched Study with Competing Risk Analysis": NA

| Diabetes Mellitus | 250 250.01 250.02 250.03 250.1 250.11 250.12 250.13 250.2 250.21 250.22 250.23 250.3 250.31 250.32 250.33 250.4 250.41 250.42 250.43 250.5 250.51 250.52 250.53 250.6 250.61 250.62 250.63 250.7 250.71 250.72 250.73 250.8 250.81 250.82 250.83 250.9 250.91 250.92 250.93 |
| --- | --- |
| Diabetic Nephropathy | 582 582.1 582.2 582.4 582.8 583.1 583.2 583.4 583.6 583.7 585 585.1 585.6 585.9 586 588 588 588.1 588.8 588.81 582.81 582.89 582.9 583 583 585.2 585.3 585.4 585.5 588.89 588.9 |
| Diabetic Retinopathy | 250.5 |
| Diabetic Neuropathy | 250.6 |
| Acute Myocardial Infarction | 410 410.01 410.02 410.1 410.11 410.12 410.2 410.21 410.22 410.3 410.31 410.32 410.4 410.41 410.42 410.5 410.51 410.52 410.6 410.61 410.62 410.7 410.71 410.72 410.8 410.81 410.82 410.9 410.91 410.92 |
| Heart Failure | 428 428.1 428.2 428.2 428.21 428.22 428.23 428.3 428.3 428.31 428.32 428.33 428.4 428.4 428.41 428.42 428.43 428.9 398.91 402.01 402.11 402.91 404.01 404.03 404.11 404.13 404.91 404.93 |
| Atrial Fibrillation | 427.31 429.4 |
| Stroke/ Transient Ischemic Attack | 435 435.1 435.2 435.3 435.8 435.9 433.81 433.91 434 436 437 437.1 433.31 433.01 434.01 434.1 434.11 434.9 434.91 437.2 437.3 437.4 437.5 437.6 437.7 437.8 437.9 430 431 432 432.1 432.9 |
| Cancer | 140.00-209.99 |
| Hypertension | 401 401.1 401.9 402 402.01 402.1 402.11 402.9 402.91 403 403.01 403.1 403.11 403.9 403.91 404404.01 404.02 404.03 404.1 404.11 404.12 404.13 404.9 404.91 404.92 404.93 405405.01 405.09 405.1 405.11 405.19 405.9 405.91 405.99 437.2 |
| Immune Mediated Colitis | 555-558 |

Supplementary table 1: ICD9 Codes for Comorbidities

|  | SGLT2i Users  (n= 18 741) | DPP4i Users  (n= 33 839) | SMD |
| --- | --- | --- | --- |
| Demographic | | | |
| Male | 10926 (58.3) | 17393 (51.4) | 0.138 |
| Age | 58.4 (11.0) | 66.9 (12.3) | 0.730 |
| Follow-up Duration | 1811 (103) | 1725 (280) |  |
| Charlson’s Score | 1.63 (1.25) | 2.54 (1.60) | 0.638 |
| Microvascular Complications | | | |
| Diabetic Retinopathy | 1981 (10.6) | 1153 (3.41) | 0.284 |
| Diabetic Nephropathy | 11965 (63.8) | 25869 (76.5) | 0.279 |
| Diabetic Neuropathy | 462 (2.46) | 327 (0.967) | 0.116 |
| Macrovascular Complications | | | |
| Heart Failure | 533 (1.67) | 1488 (4.40) | 0.083 |
| Acute Myocardial Infarct | 703 (3.75) | 952 (2.81) | 0.053 |
| Stroke/ Transient Ischemic Attack | 564 (3.01) | 1454 (4.30) | 0.069 |
| Atrial Fibrillation | 472 (2.52) | 1127 (3.33) | 0.048 |
| Other Comorbidities | | | |
| Hyperlipidemia | 774 (4.13) | 888 (2.62) | 0.083 |
| Hypertension | 5054 (27.0) | 9517 (28.1) | 0.026 |
| Immune Mediated Enterocolitis | 1089 (5.80) | 1953 (5.78) | 0.002 |
| Other Cancer | 347 (1.85) | 1195 (3.53) | 0.104 |
| Anti-Diabetic and Lipid-Lowering Medications | | | |
| Metformin | 17585 (93.8) | 29627 (87.6) | 0.215 |
| Sulphonylurea | 13651 (72.8) | 27838 (82.3) | 0.228 |
| Insulin | 10075 (53.7) | 17508 (51.7) | 0.040 |
| Acarbose | 856 (4.57) | 593 (1.75) | 0.161 |
| Thiazolidinedione | 5498 (29.3) | 4861(14.4) | 0.368 |
| Glucagon-Like-Peptide Agonist | 1448 (7.72) | 149 (4.40) | 0.374 |
| Statins | 15493 (82.6) | 10881(32.2) | 1.187 |
| Complete Blood Count | | | |
| Red Cell Count | 4.74 (0.588) | 4.35 (0.705) | 0.603 |
| Mean Corpuscular Volume | 86.7 (7.15) | 87.6 (7.75) | 0.126 |
| Mean Corpuscular Hemoglobin Concentration | 29.2 (2.86) | 29.6 (3.09) | 0.149 |
| White Cell Count | 7.98 (2.60) | 8.04 (3.21) | 0.022 |
| Neutrophil | 5.13 (2.41) | 5.49 (3.04) | 0.130 |
| Lymphocyte | 2.16 (0.907) | 1.88 (0.898) | 0.309 |
| Neutrophil to Lymphocyte Ratio | 2.96 (3.77) | 4.01 (5.34) | 0.228 |
| Eosinophil | 0.216 (0.193) | 0.220 (0.282) | 0.012 |
| Platelet | 245 (67.6) | 236 (75.1) | 0.118 |
| Platelet to Lymphocyte Ratio | 133 (157) | 155 (146) | 0.143 |
| Renal Function Test | | | |
| Sodium | 139 (2.74) | 139 (3.12) | 0.045 |
| Potassium | 4.31 (0.434) | 4.37 (0.504) | 0.140 |
| Urea | 5.73 (2.03) | 7.21 (4.20) | 0.448 |
| Creatinine | 78.8 (28.3) | 107 (96.1) | 0.395 |
| Estimated Glomerular Filtration Rate | 84.4 (22.4) | 68.4 (27.6) | 0.635 |
| Liver Function Test | | | |
| Alkaline Phosphatase | 73.8 (26.1) | 79.6 (36.7) | 0.184 |
| Aspartate Transaminase | 28.2 (28.0) | 28.0 (65.1) | 0.004 |
| Alanine Transaminase | 32.2 (29.7) | 26.5 (37.0) | 0.171 |
| Bilirubin | 11.4 (6.16) | 11.0 (7.59) | 0.054 |
| Protein | 74.4 (4.94) | 73.4 (5.92) | 0.179 |
| Albumin | 42.5 (3.27) | 40.9 (4.34) | 0.406 |
| Lipid Profile | | | |
| Triglyceride | 1.80 (1.72) | 1.65 (1.31) | 0.097 |
| Total Cholesterol | 4.32 (1.01) | 4.32 (0.978) | 0.002 |
| High-Density Lipoprotein | 1.16 (0.309) | 1.22 (0.342) | 0.160 |
| Low-Density Lipoprotein | 2.38 (0.804) | 2.38 (0.803) | 0.001 |
| Glucose Control | | | |
| HbA1c | 8.03 (1.67) | 8.08 (1.69) | 0.031 |
| Fasting Blood Glucose | 9.21 (3.65) | 8.74 (4.05) | 0.122 |
| Outcomes | | | |
| Death | 488 (2.60) | 5417 (16.0) |  |
| Incident CRC | 94 (0.501) | 377 (1.11) |  |

Supplementary Table 2: Baseline Demographics of SGLT2I and DPP4I Users Before Propensity Score Matching

| Variable | Hazard Ratio | 95% CI | P-value |
| --- | --- | --- | --- |
| Demographic |  |  |  |
| Male | 1.24 | 0.920-1.67 | 0.158 |
| Age | 1.06 | 1.04-1.07 | <0.001 |
| Charlson’s Score | 1.40 | 1.29-1.51 | <0.001 |
| Microvascular Complications |  |  |  |
| Diabetic Retinopathy | 0.854 | 0.437-1.67 | 0.645 |
| Diabetic Nephropathy | 1.71 | 1.22-2.4 | 0.002 |
| Chronic Kidney Disease Stage | 1.26 | 1.04-1.52 | 0.018 |
| Diabetic Neuropathy | 0.725 | 0.180-2.92 | 0.651 |
| Macrovascular Complications |  |  |  |
| Heart Failure | 2.20 | 1.16-4.17 | 0.015 |
| Acute Myocardial Infarction | 1.83 | 0.935-3.57 | 0.078 |
| Stroke/ Transient Ischemic Attack | 1.26 | 0.594-2.69 | 0.544 |
| Atrial Fibrillation | 0.898 | 0.334-2.42 | 0.832 |
| Other Comorbidities |  |  |  |
| Hyperlipidemia | 0.755 | 0.280-2.03 | 0.579 |
| Hypertension | 1.31 | 0.955-1.81 | 0.094 |
| Immune Mediated Enterocolitis | 0.960 | 0.491-1.88 | 0.906 |
| Other Cancer | 3.12 | 1.74-5.59 | <0.001 |
| Anti-Diabetic and Lipid-Lowering Medications | |  |  |
| Metformin | 0.635 | 0.390-1.03 | 0.067 |
| Sulphonylurea | 1.08 | 0.746-1.57 | 0.677 |
| Insulin | 2.36 | 1.74-3.21 | <0.001 |
| Acarbose | 1.42 | 0.666-3.02 | 0.365 |
| Thiazolidinedione | 0.447 | 0.278-0.718 | <0.001 |
| Glucagon-Like-Peptide Agonist | 0.355 | 0.050-2.53 | 0.302 |
| Statins | 1.08 | 0.767-1.52 | 0.661 |
| Complete Blood Count |  |  |  |
| Red Cell Count | 0.580 | 0.431-0.781 | <0.001 |
| Mean Corpuscular Volume | 1.01 | 0.980-1.03 | 0.637 |
| Mean Corpuscular Hemoglobin Concentration | 0.992 | 0.932-1.06 | 0.798 |
| White Cell Count | 1.02 | 0.959-1.09 | 0.492 |
| Neutrophil | 1.05 | 0.987-1.13 | 0.115 |
| Lymphocytes | 0.717 | 0.530-0.969 | 0.030 |
| Neutrophil to Lymphocyte Ratio | 1.02 | 0.982-1.06 | 0.312 |
| Eosinophil | 0.812 | 0.250-2.63 | 0.728 |
| Platelet | 1.000 | 0.997-1.00 | 0.745 |
| Platelet to Lymphocyte Ratio | 1.00 | 0.999-1.00 | 0.880 |
| Renal Function Test |  |  |  |
| Sodium | 1.03 | 0.979-1.09 | 0.235 |
| Potassium | 1.33 | 0.975-1.82 | 0.072 |
| Urea | 1.04 | 1.00-1.08 | 0.029 |
| Creatinine | 1.00 | 0.999-1.00 | 0.330 |
| Estimated Glomerular Filtration Rate | 0.987 | 0.981-0.993 | <0.001 |
| Liver Function Test |  |  |  |
| Alkaline Phosphatase | 1.00 | 0.998-1.01 | 0.230 |
| Aspartate Transaminase | 1.000 | 0.991-1.01 | 0.959 |
| Alanine Transaminase | 0.996 | 0.988-1.00 | 0.385 |
| Bilirubin | 0.992 | 0.962-1.02 | 0.595 |
| Protein | 0.989 | 0.957-1.02 | 0.506 |
| Albumin | 0.965 | 0.925-1.01 | 0.101 |
| Lipid Profile |  |  |  |
| Triglyceride | 0.932 | 0.810-1.07 | 0.322 |
| Total Cholesterol | 0.941 | 0.800-1.11 | 0.460 |
| High-Density Lipoprotein | 0.965 | 0.598-1.56 | 0.883 |
| Low-Density Lipoprotein | 0.961 | 0.790-1.17 | 0.695 |
| Glucose Control |  |  |  |
| Glycated Hemoglobin | 0.989 | 0.907-1.08 | 0.795 |
| Fasting Blood Glucose | 1.01 | 0.967-1.05 | 0.778 |

Supplementary Table 3: Univariable Cause-Specific Cox Regression after Propensity Score Matching

| Variable | Hazard Ratio | | | 95% CI | P-value | |
| --- | --- | --- | --- | --- | --- | --- |
| Demographic |  | | |  |  | |
| Male | 1.34 | | | 1.11-1.61 | 0.002 | |
| Age | 1.05 | | | 1.04-1.06 | <0.001 | |
| Charlson’s Score | 1.34 | | | 1.28-1.40 | <0.001 | |
| Microvascular Complications |  | | |  |  | |
| Diabetic Retinopathy | 0.854 | | | 0.437-1.67 | 0.645 | |
| Diabetic Nephropathy | 1.77 | | | 1.40-2.24 | <0.001 | |
| Stage of Diabetic Nephropathy | 1.22 | | | 1.11-1.34 | <0.001 | |
| Diabetic Neuropathy | 1.71 | | | 1.22-2.40 | 0.002 | |
| Macrovascular Complications |  | | |  |  | |
| Heart Failure | 1.28 | | | 0.823-1.98 | 0.276 | |
| Acute Myocardial Infarction | 1.83 | | | 0.935-3.57 | 0.078 | |
| Acute Myocardial Infarction | 1.63 | | | 1.07-2.48 | 0.022 | |
| Atrial Fibrillation | 1.11 | | | 0.666-1.86 | 0.682 | |
| Other Comorbidities |  | | |  |  | |
| Hyperlipidemia | 0.755 | | | 0.280-2.03 | 0.579 | |
| Hypertension | 1.30 | | | 1.07-1.57 | 0.008 | |
| Immune Mediated Enterocolitis | 0.785 | | | 0.517-1.19 | 0.255 | |
| Other Cancer | 4.09 | | | 3.04-5.50 | <0.001 | |
| Anti-diabetic and Lipid-lowering Medications |  | | |  |  | |
| Metformin | 0.635 | | | 0.390-1.03 | 0.067 | |
| Sulphonylurea | 1.28 | | | 1.01-1.62 | 0.042 | |
| Insulin | 2.35 | | | 1.93-2.87 | <0.001 | |
| Acarbose | 0.417 | | | 0.666-3.02 | 0.365 | |
| Thiazolidinedione | 0.447 | | | 0.278-0.718 | 0.001 | |
| Glucagon-Like-Peptide Agonist | 0.355 | | | 0.050-2.53 | 0.302 | |
| Statins | 1.08 | | | 0.767-1.52 | 0.661 | |
| Complete Blood Count |  | | |  |  | |
| Red Cell Count | 0.664 | | | 0.559-0.787 | <0.001 | |
| Mean Corpuscular Volume | 1.01 | | | 0.980-1.033 | 0.637 | |
| Mean Corpuscular Hemoglobin Concentration | 0.992 | | | 0.932-1.06 | 0.798 | |
| White Cell Count | 1.01 | | | 0.989-1.04 | 0.274 | |
| Neutrophil | 1.04 | | | 1.00-1.08 | 0.049 | |
| Lymphocyte | 0.717 | | | 0.530-0.969 | 0.030 | |
| Neutrophil to Lymphocyte Ratios | 1.02 | | | 1.01-1.04 | 0.005 | |
| Eosinophil | 1.16 | | | 0.849-1.59 | 0.352 | |
| Platelet | 1.00 | | | 0.999-1.00 | 0.749 | |
| Platelet to Lymphocyte Ratios | 1.00 | | | 1.00-1.00 | 0.031 | |
| Renal Function Test |  | | |  |  | |
| Sodium | 1.02 | | | 0.992-1.06 | 0.148 | |
| Potassium | 1.09 | | | 0.902-1.32 | 0.370 | |
| Urea | 1.03 | | | 1.01-1.05 | 0.007 | |
| Creatinine | 1.00 | | | 1.00-1.00 | 0.047 | |
| Estimated Glomerular Filtration Rate | 0.987 | | | 0.981-0.993 | <0.001 | |
| Liver Function Test |  | | |  |  | |
| Alkaline Phosphatase | 1.00 | | | 1.00-1.00 | 0.113 | |
| Aspartate Transaminase | 1.00 | | | 0.991-1.01 | 0.959 | |
| Alanine Transaminase | 0.996 | | | 0.988-1.00 | 0.385 | |
| Bilirubin | 0.992 | | | 0.962-1.02 | 0.595 | |
| Protein | 0.989 | | | 0.957-1.02 | 0.506 | |
| Albumin | 0.965 | | | 0.925-1.01 | 0.101 | |
| Lipid Profile | |  |  | | |  |
| Triglyceride | | 0.932 | 0.810-1.07 | | | 0.322 |
| Total Cholesterol | | 0.900 | 0.813-1.00 | | | 0.043 |
| High-Density Lipoprotein | | 0.965 | 0.598-1.56 | | | 0.883 |
| Low-Density Lipoprotein | | 0.961 | 0.790-1.17 | | | 0.695 |
| Glucose Control | |  |  | | |  |
| Glycated Hemoglobin | | 0.989 | 0.907-1.08 | | | 0.795 |
| Fasting Blood Glucose | | 1.01 | 0.967-1.05 | | | 0.778 |

Supplementary Table 4: Univariable Cause-Specific Cox Regression Before Propensity Score Matching

| Variables | Hazard ratios | 95% CI | P-value |
| --- | --- | --- | --- |
| Before Propensity Score Matching | | | |
| Model 1 | 0.446 | 0.355-0.558 | <0.001 |
| Model 2 | 0.623 | 0.289-0.794 | <0.001 |
| Model 3 | 0.622 | 0.488-0.793 | <0.001 |
| Model 4 | 0.615 | 0.481-0.788 | <0.001 |
| Model 5 | 0.625 | 0.487-0.801 | <0.001 |
| Model 6 | 0.608 | 0.470-0.787 | <0.001 |
| After Propensity Score Matching | | | |
| Model 1 | 0.543 | 0.401-0.734 | <0.001 |
| Model 2 | 0.570 | 0.420-0.773 | <0.001 |
| Model 3 | 0.566 | 0.417-0.769 | <0.001 |
| Model 4 | 0.566 | 0.417-0.769 | <0.001 |
| Model 5 | 0.575 | 0.368-0.896 | 0.015 |
| Model 6 | 0.552 | 0.400-0.762 | <0.001 |

Supplementary Table 5: Univariable and Multivariable Fine-Gray’s Subdistribution Regression before and after Propensity Score Matching

Model 1: Unadjusted hazard ratio. Model 2: Adjusted for age and gender. Model 3: Adjusted for age, gender and HbA1c. Model 4: Adjusted for age, gender, HbA1c and use of other medications. Model 5: Adjusted for age, gender, HbA1c, use of medications and comorbidities. Model 6: Adjusted for age, gender, HbA1c, use of medications, comorbidities and HDL.

|  | Hazard ratios | 95% CI | P-value |
| --- | --- | --- | --- |
| Male | | | |
| Model 1 | 0.478 | 0.322-0.709 | <0.001 |
| Model 2 | 0.501 | 0.337-0.746 | <0.001 |
| Model 3 | 0.500 | 0.336-0.744 | <0.001 |
| Model 4 | 0.506 | 0.340-0.754 | <0.001 |
| Model 5 | 0.505 | 0.339-0.752 | <0.001 |
| Model 6 | 0.484 | 0.318-0.736 | <0.001 |
| Female | | | |
| Model 1 | 0.651 | 0.406-1.04 | 0.075 |
| Model 2 | 0.694 | 0.432-1.11 | 0.130 |
| Model 3 | 0.695 | 0.433-1.12 | 0.130 |
| Model 4 | 0.658 | 0.405-1.07 | 0.091 |
| Model 5 | 0.657 | 0.404-1.07 | 0.091 |
| Model 6 | 0.642 | 0.386-1.07 | 0.089 |

Supplementary Table 6: Univariable and Multivariable Fine-and-Gray Subdistribution Regression after Propensity Score Matching in Patient Subgroup Stratified by Gender

Model 1: Unadjusted hazard ratio. Model 2: Adjusted for age. Model 3: Adjusted for age and HbA1c. Model 4: Adjusted for age, HbA1c and use of other medications. Model 5: Adjusted for age, HbA1c, use of medications and comorbidities. Model 6: Adjusted for age, gender, HbA1c, use of medications, comorbidities, and HDL.

|  | Hazard ratios | 95% CI | P-value |
| --- | --- | --- | --- |
| Subjects < 65 Years Old | | | |
| Model 1 | 0.349 | 0.212-0.575 | <0.001 |
| Model 2 | 0.344 | 0.208-0.567 | <0.001 |
| Model 3 | 0.344 | 0.208-0.567 | <0.001 |
| Model 4 | 0.326 | 0.197-0.541 | <0.001 |
| Model 5 | 0.328 | 0.198-0.546 | <0.001 |
| Model 6 | 0.300 | 0.176-0.511 | <0.001 |
| Subject ≥ 65 Years Old | | | |
| Model 1 | 0.766 | 0.521-1.13 | 0.180 |
| Model 2 | 0.782 | 0.529-1.15 | 0.220 |
| Model 3 | 0.780 | 0.528-1.15 | 0.201 |
| Model 4 | 0.791 | 0.535-1.17 | 0.240 |
| Model 5 | 0.783 | 0.528-1.16 | 0.220 |
| Model 6 | 0.822 | 0.540-1.25 | 0.360 |

Supplementary Table 7: Univariable and Multivariable Fine-and-Gray Subdistribution Regression after Propensity Score Matching in Patient Subgroup Stratified by Age

Model 1: Unadjusted hazard ratio. Model 2: Adjusted for gender. Model 3: Adjusted for gender and HbA1c. Model 4: Adjusted for gender, HbA1c and use of other medications. Model 5: Adjusted for gender, HbA1c, use of medications and comorbidities. Model 6: Adjusted for age, gender, HbA1c, use of medications, comorbidities, and HDL.

|  | Hazard ratios | 95% CI | P-value |
| --- | --- | --- | --- |
| eGFR ≥ 45 mL/min/ 1.73m^2^ | | | |
| Model 1 | 0.492 | 0.374-0.648 | <0.001 |
| Model 2 | 0.624 | 0.464-0.840 | 0.002 |
| Model 3 | 0.624 | 0.464-0.839 | 0.002 |
| Model 4 | 0.608 | 0.450-0.823 | 0.001 |
| Model 5 | 0.620 | 0.458-0.839 | 0.002 |
| Model 6 | 0.593 | 0.434-0.811 | 0.001 |
| eGFR < 45 mL/min/ 1.73m^2^ | | | |
| Model 1 | 0.546 | 0.201-1.48 | 0.230 |
| Model 2 | 0.616 | 0.226-1.68 | 0.340 |
| Model 3 | 0.617 | 0.226-1.68 | 0.350 |
| Model 4 | 0.609 | 0.222-1.67 | 0.330 |
| Model 5 | 0.620 | 0.227-1.69 | 0.350 |
| Model 6 | 0.678 | 0.247-1.86 | 0.450 |

Supplementary Table 8: Univariable and Multivariable Fine Gray’s Subdistribution Regression before Propensity Score Matching in Patient Subgroup Stratified by eGFR

Model 1: Unadjusted hazard ratio. Model 2: Adjusted for age and gender. Model 3: Adjusted for age, gender and HbA1c. Model 4: Adjusted for age, gender, HbA1c and use of other medications. Model 5: Adjusted for age, gender, HbA1c, use of medications and comorbidities. Model 6: Adjusted for age, gender, HbA1c, use of medications, comorbidities and HDL.

|  | Hazard ratios | 95% CI | P-value |
| --- | --- | --- | --- |
| Model 1 | 0.389 | 0.092-1.64 | 0.197 |
| Model 2 | 0.405 | 0.097-1.68 | 0.213 |
| Model 3 | 0.374 | 0.090-1.55 | 0.176 |
| Model 4 | 0.332 | 0.078-1.41 | 0.135 |
| Model 5 | 0.285 | 0.066-1.25 | 0.097 |

Supplementary Table 9: Univariable and Multivariable Cox Regression Before Propensity Score Matching for All-Cause Mortality in Patients with Incident CRC

Model 1: Unadjusted hazard ratio. Model 2: Adjusted for age and gender. Model 3: Adjusted for age, gender and HbA1c. Model 4: Adjusted for age, gender, HbA1c and use of other medications. Model 5: Adjusted for age, gender, HbA1c, use of medications and comorbidities.
